# Barriers and facilitators for the implementation of wiki- and blog-based Virtual Learning Environments as tools for improving collaborative learning in the Bachelor of Nursing degree

**DOI:** 10.1101/2024.09.16.24313727

**Authors:** Beatriz Rodríguez-Martín, Rubén Mirón-González, María del Carmen Zabala-Baños, Montserrat Pulido-Fuentes, Esmeralda Santacruz-Salas, Carlos A. Castillo-Sarmiento

## Abstract

There is little research on teachers’ perceptions of the use of Virtual Learning Environments (VLEs) in undergraduate nursing education. The aim of this study was to understand teachers’ perceptions of the barriers and facilitators for the implementation of wiki- and blog-based VLEs as tools to improve collaborative learning.

A qualitative action-research study in an intentional opinion-intensity sample of teachers on the Bachelor of Nursing degree course was used. The data were collected and analysed according to the SWOT approach, the constant comparison method, and the coding process.

The analysis highlighted weaknesses (e.g., limited audio-visual communication and feedback, privacy concerns), threats (e.g., training, pre-planning, and time cost), strengths (e.g., being intuitive, friendly and accessible, creating shared knowledge) and opportunities (e.g., improving technological skills, teaching innovation, evaluation and peer learning) for the incorporation of VLEs to promote collaborative learning in nursing training.

Although both our study and the existing literature on the subject show many advantages that encourage the implementation of VLEs in nursing training, the barriers described in this paper cannot be ignored in the context in which the university currently finds itself.

These barriers and facilitators to implementing VLEs in nursing training will be of interest to educational authorities.

## Main text introduction

According to the UK’s Joint Information Systems Committee, the term Virtual Learning Environments (VLEs) refers to the components in which learners and tutors participate in various kinds of ’online’ interaction, including online learning. Since the end of the 20th century, Faculties and Centres of Health Sciences have integrated VLEs into their undergraduate training through computer aids and web-based learning programmes (Barrios Araya et al., 2011). Since its inception, the field of nursing education has been committed to using VLEs as an asset in distance training due to the benefits it provides in the teaching–learning–evaluation process (Cruz, 2009; Holanda et al., 2013).

Among the best known, used and developed VLEs are Moodle, WebCT and Blackboard. These platforms are characterised by the integration of web tools such as e-mail, discussion forums, wikis and blogs. Wikis and blogs allow people to interact and communicate when creating shared documents, and offer a multitude of possibilities in health sciences and nursing education (González Hernando et al., 2015; Rasmussen et al., 2013; Rodríguez Martín, 2020). Both tools are used as shared spaces which increase students’ motivation, satisfaction and performance, provide pathways to give feedback, favour the development of students’ critical and creative thinking (García-Martín & García-Sánchez, 2015) and capability for self-reflection, thus increasing the quality of their learning (Reed & Edmunds, 2015).

A VLE facilitates the development of active methodologies such as Problem Based Learning (PBL), Project Based Learning, Flipped Classroom or Peer Instruction and Team Based Learning (Fonseca & Mattar, 2017). Among these active methodologies is collaborative learning, which has been applied in the training of nurses for more than two decades (Zhang & Cui, 2018) and has become a popular methodology in conjunction with VLEs due to the characteristics of globality, interactivity and immediacy offered by the latter (Zañartu Correa, 2003). Collaborative learning fosters a positive attitude to learning (Wong, 2017) and improves learning initiative, cooperative awareness, critical thinking and communication skills (Baghcheghi et al., 2011; Lee et al., 2016; Papathanasiou et al., 2014). The approach is a recommended methodology in theoretical and practical nursing training (Zhang & Cui, 2018) because of its ability to promote cognitive integration and basic skills in health science professionals (Ignacio & Chen, 2020).

Furthermore, VLE-based collaborative learning entails a shift in the role of the teacher, who must now act as a guide to learning. Moreover, teachers need to be familiar with and know how to administer VLEs, but also be capable of motivating and guiding students to help them manage the emotions that may arise in the face of conflicts in the context of collaborative learning (Martínez Noris & Ávila Aguilera, 2014). However, in terms of the attitudes of teachers toward the implementation of collaborative learning through VLEs, there is little research on teachers’ perceptions of the use of VLEs in undergraduate nursing education (Lai & Bower, 2020; Lin et al., 2015; Moule et al., 2011; Souza Leite et al., 2016) and, to our knowledge, no research on the application of VLEs in collaborative learning. Such applications are especially relevant in the current global situation, where VLEs have not just proven useful, but essential, tools for completing the training of nurses during the COVID-19 pandemic (Jackson et al., 2020; Ng & Or, 2020).

The objective of this study was to understand teachers’ perceptions of the barriers and facilitators for the implementation of wiki- and blog-based VLEs as tools to improve collaborative learning.

## Materials and methods

Qualitative study based on the action-research approach, within the framework of a broader research project carried out during the 2018-19 academic year with the aim of training students and teaching staff of the Bachelor of Nursing degree at the University of Castilla-La Mancha (UCLM, Spain) to use wiki- and blog-based VLEs intended to enhance teaching–learning–evaluation processes. The action-research paradigm was chosen for its usefulness in improving teaching through change, allowing for systematic reflection on the study phenomenon in order to optimise teaching–learning–evaluation processes (Bausela Herreras, 2004).

According to the principles of action research, the research project followed a spiral model based on cycles that included planning (analysing the situation, developing an action plan to train both students and teachers in the use of VLEs), action (implementation of VLEs in the individual subjects), observation, and reflection or evaluation (Bausela Herreras, 2004).

The project involved seven full-time instructors of the Bachelor of Nursing degree course in the Faculty of Health Sciences (FHS) and Faculty of Physiotherapy and Nursing (FPN) at UCLM, along with all students of the ten participating Bachelor of Nursing modules: Fundamentals of Nursing II, Psychology, Introduction to Public Health, and Nutrition and Dietetics in the first academic year; Health Psychology, Medical-Surgical Nursing III, and Clinical Stays I in the second year; Geriatric Nursing, Community Nursing II, and Medical-Surgical Nursing IV in the third year; and Practicum II in the fourth year (a total of 328 students: 65 and 98 students in the first years of FHS and FPN, respectively, 65 students in the second year of FHS, 65 in the third year of FHS and 50 in the fourth year of FHS).

a. Participants and recruitment

The qualitative study used an intentional opinion-intensity sample, which included seven full-time instructors from the two faculties teaching the ten modules covered by the project. The following inclusion criteria were used for the selection of the participants: 1) to be a lecturer on the Bachelor of Nursing degree in the FHS or FPN, 2) to be teaching one of the subjects covered by the project, 3) to have used a wiki- or blog-based VLE in that subject, and 4) to agree to participate voluntarily in the research. The exclusion criterion employed was to be a part-time teacher or not to be responsible for the subject.

b. Data collection and analysis

Throughout the 2018-19 academic year, the teaching staff involved in the project engaged in a process of ongoing observation and reflection to collect relevant information covering their experiences of the whole process of implementing the VLE in their subject, paying special attention to the barriers and facilitators for implementation, student participation, and any suggestions for improvement. Once each course element involving a VLE had been completed, the respective teaching staff were asked to carry out a SWOT (Strengths, Weaknesses, Opportunities and Threats) analysis of the process of implementing the VLE in their subject, which included internal (strengths and weaknesses) and external (opportunities and threats) analyses.

Once the SWOT assessment had been completed, the data were anonymised for further analysis. Two researchers (BRM and CACS) conducted independent discourse analyses of each SWOT, subsequently discussing their results to reach a consensus.

The data analysis applied the constant comparison method and the open, axial and selective coding processes (Rodríguez-Martín et al., 2013; Silverman, 2016). Data saturation was reached during the analysis process.

c. Ethical considerations

The study complied with fundamental ethical principles and current legislation. The participants’ data were anonymised before being analysed. All participants freely agreed to participate in the study and signed the informed consent declaration. Due to the nature of the study, approval by the Research Ethics Committee was not required.

## Results

As part of the research, VLEs based on blogs and wikis were implemented in the participating subjects, with the majority choosing to use wikis. The support platform for the wikis was the UCLM Virtual Campus, a Moodle-based learning management system, while the blogs were hosted on WordPress.

To present the results more clearly, the barriers and facilitators for the implementation of wiki- and blog-based VLEs are discussed separately. Six of the instructors (accounting for eight of the subjects studied) designed and implemented wiki-based VLEs, while one instructor used blogs in two subjects.

### Barriers and facilitators for the implementation of wiki-based VLEs

On one hand, the main weakness of the Moodle wiki identified by the teaching staff was that the tool did not allow complex editing of data or the export of complete data. In addition, the limited options for audio-visual communication and maintaining privacy were cited as other weaknesses of the wiki. On the other hand, although the wiki did allow some feedback to be provided to students, the evaluation and feedback possibilities with this tool were found somewhat lacking. Finally, the need to pre-register using the Virtual Campus user and password in order to access the wiki was considered another weakness.

In terms of threats, teachers felt that both students and teachers should be given training to use the wiki before proposing that students undertake activities using it. It was also necessary for teachers to plan the activities, learning objectives and mode of evaluation beforehand. The need to provide feedback to students, and the associated time cost, were other threats cited by teachers.

According to the teachers, the main strengths of the wiki for use in collaborative learning were that it was free, intuitive, and user-friendly, and that it was integrated into the institutional learning management system. This not only made it an accessible and convenient tool for students, but also meant technical support was available from the institutional information technology service. Additional positive aspects highlighted by the participants included the accessibility of the tool, the possibility of creating shared knowledge, the ability to create and immediately disseminate knowledge (enabling students to visualise the work of the whole class), the promotion of good organisation of content, and the support for the insertion of videos and documents in PDF, Word, Excel or other file formats.

The teachers also highlighted several opportunities for the use of wikis for the collaborative learning. One main advantage was that the use of these tools was considered to offer an opportunity to improve technological skills and modernise the ways in which students and teachers communicate and interact. In addition, wikis are one means to design more accessible strategies to encourage learning participation and promote innovation in teaching. Furthermore, teachers considered that wikis promoted cooperative learning, allowed the material created by each group to be available to all students, encouraged students to learn from each other’s work, favoured peer assessment, and promoted continuous learning. The teachers also evaluated very positively the opportunity provided by the wikis for the same case to be worked on in parallel across several subjects, avoiding saturating the students with multiple assignments for several subjects. Finally, the case studies helped students to develop a comprehensive view of the health problems that might affect a person.

### Barriers and facilitators for the implementation of VLEs based on blogs

The participants found the main weakness of blogs to be their complexity, considering that a priori they could be difficult for students and teachers to use.

In terms of threats, the teachers stressed that the tool required previous training, and also highlighted the students’ responses to using blogs if they thought that they might not use them again after the course was over. Furthermore, teachers stated that it was not easy to design teaching activities using such an environment.

The main perceived strengths of blogs were the online nature of the tool, and the fact that they are asynchronous, free and easily linked to social networks. Finally, among the opportunities for using blogs, teachers highlighted the fact that both healthcare professionals and scientific disseminators frequently use them as a tool, that blogs are versatile, and that they can incorporate many kinds of multimedia material.

## Discussion

This analysis of the implementation of VLEs in teaching in nursing highlights a significant complexity that must be considered when incorporating such tools into university teaching. Furthermore, our findings echo many of the positive factors recognised in the literature for these types of environment (Kala et al., 2010), such as free access and the possibility of working asynchronously. Similarly, the use of these environments is perceived by teaching staff to be a tool that improves students’ ICT skills, but also encourages other skills not necessarily linked to the use of technology, such as better cooperation when working in groups and improved integration of acquired knowledge.

However, negative aspects have also been highlighted, mainly the need to invest resources in the training of students and teachers in order to impart an adequate understanding of these tools, enabling their full potential to be exploited.

This need to invest resources in training users cannot be neglected in the analysis. In fact, appropriate academic literacy, in a broader sense than just electronic learning methods, is considered fundamental for nursing students, (Jefferies et al., 2018). In the current context, with public universities having less resources available to them, both financially and in terms of teaching time, while the number of students per classroom is increasing and the quality criteria for obtaining certifications are becoming more stringent in a highly competitive environment, introducing a methodology that requires additional resources for its implementation does not seem a simple task. Moreover, we note that that the use of VLEs, although potentially providing pedagogical benefits for student training, brings with it a significant increase in the teacher’s workload (Bai et al., 2016) that institutions should take into account. We must further remember, however, that a possible publication bias toward reporting successful experiences using technology in education has already been identified in the literature (Conde-Caballero et al., 2019).

The use of tools that are not freeware or that, although freeware, are hosted on servers outside the university, raises some ethical aspects that should also be highlighted. For instance, undertaking activities on platforms outside the university itself implies a need for students to register on these platforms and accept the respective terms of service, which are not always transparent and can change depending on the interests of the company providing the service. In addition, tools hosted on external servers may be subject to attacks that can endanger or expose both the content created and the personal data in users’ accounts. Finally, the fact that the use of digital technology in higher education represents an enormous commercial interest for technology companies, often without pedagogical justification, should not be disregarded (Bartolomé et al., 2018), as this factor may exert some influence on purely academic aspects.

Although it has been previously mentioned in the literature as a barrier to the implementation of VLEs (Chawinga, 2017), the cost or quality of the network or the lack of hardware allowing access to the technology does not seem to be a significant barrier in the context we studied.

Our analysis does suggest that teaching staff perceive there to be potentially attractive benefits for university students, especially those of nursing, in three particular aspects: teamwork, use of ICT, and integration of acquired knowledge. Despite the considerations outlined above, these technologies have the potential to improve the way nurses work as a team, which is one of the pillars of professional nursing practice (Barton et al., 2018). Although there is no single definition of teamwork that all health professionals agree upon (Xyrichis & Ream, 2008), improved communication between members of the healthcare team is key to better teamwork (Dingley et al., 2008). One way to achieve this is through the appropriate use of technology (Wickersham et al., 2018). Furthermore, as already described in the literature, the use of ICT could improve nursing practice by enhancing the planning, organisation and documentation of care, among other indicators (Cherrill & Linsley, 2017; Rouleau et al., 2017). This would ultimately translate into improved quality and safety of care provided.

However, despite the widespread appreciation of the potential of incorporating ICT into nursing training, institutions are not providing sufficient support for the integration of these technologies (Button et al., 2014; Nwozichi et al., 2019). Finally, the effect of enhancing the integration of acquired knowledge and developing critical thinking on improving care has been explored previously (Reale et al., 2018). These are universal goals in university institutions, although they are particularly important skills in the field of health sciences given the speed with which scientific knowledge is revised and updated. Therefore, if nurses acquire the necessary skills to integrate new knowledge during their training, they will be more aware of the changes that occur in nursing practice and, as a result, better equipped with the life-long learning skills required to provide safe, evidence-based care.

Regarding the use of VLEs in learning, that is, whether these tools are better or worse than others for learning, we echo the ideas put forward by Biesta (Biesta, 2009), which continue to apply today. Although the use of technologies in teaching environments is usually discussed in terms of their effect on learning, this clearly limits the ways in which digital technologies are understood, as there are many other aspects of education that go far beyond issues related to the acquisition of content (Ferrés et al., 2018), such as the aspects we have already highlighted in this work. In our opinion, placing too much focus on the link between the use of technology and the acquisition of content ignores much of the potential that VLEs bring to university teaching in terms of developing other skills and aptitudes. This is even more important considering that comparative quality studies have already shown that, in terms of learning, digital approaches are not necessarily better than traditional methods (Berg & Steinsbekk, 2020; Seifert et al., 2019). This aspect is especially relevant if, as pointed out earlier, the technology has not been developed by teachers but provided by an external company which essentially, through the way in which the tool is programmed, determines what is and is not possible in the teaching setting. In this regard, one characteristic of the present work which could be considered a limitation is that our objectives did not include the compilation of the different pedagogical foundations used by each of the teachers in their VLEs, a fundamental piece of information which would be needed to fully understand a process as complex as learning (Decuypere, 2019). Instead, the present work focused on the perception of the teachers regarding the possible implementation of the technology.

Although both our study and the previous literature on the subject show many advantages that encourage the implementation of VLEs in nursing training, the barriers described in this paper, mainly related to the resources that have to be mobilised for the correct implementation of these technologies, cannot be ignored in the situation in which universities currently finds themselves. Furthermore, we reiterate that the use of wikis has great potential as they can be integrated into the institution’s Virtual Campus, while one main strength of blogs is their connectivity to social networks. The present work provides evidence of the barriers and facilitators encountered for the implementation of VLEs in university-level nursing training, and as such may be of interest to educational authorities interested in promoting the incorporation of these tools into their teaching environments.

## Declarations

### Funding

This work was supported by the National Conference of Deans of Nursing, Spain.

## Data Availability

All data produced in the present study are available upon reasonable request to the authors.

## Acknowledgments

None.

## Declaration of interest statement

None.

## Notes

### Competing Interest Statement

The authors have declared no competing interest.

